# Adaptive, Unlabeled and Real-time Approximate-Learning Platform (AURA) for Personalized Epileptic Seizure Forecasting

**DOI:** 10.1101/2021.09.30.21264287

**Authors:** Yikai Yang, Nhan Duy Truong, Jason K. Eshraghian, Armin Nikpour, Omid Kavehei

## Abstract

A high performance event detection system is all you need for some predictive studies. Here, we present AURA: an <u>A</u>daptive forecasting model trained with <u>U</u>nlabeled, <u>R</u>eal-time data using internally generated <u>A</u>pproximate labels on-the-fly. By harnessing the correlated nature of time-series data, a pair of detection and prediction models are coupled together such that the detection model generates labels automatically, which are then used to train the prediction model. AURA relies on several simple principles and assumptions: (i) the performance of an event prediction/forecasting model in the target application remains below the performance of an event detection model, (ii) detected events are treated as weak labels and deemed reliable enough for online training of a predictive model, and (iii) system performance and/or system responsive feedback characteristics can be tuned for a subject-under-test. For example, in medical patient monitoring, this enables personalizing forecasting models. Seizure prediction is identified as an ideal test case of AURA, as pre-ictal brainwaves are patient-specific and tailoring models to individual patients can significantly improve forecasting performance. AURA is used to generate an individual forecasting model for 10 patients, showing an average relative improvement in sensitivity by 14.30% and reduction in false alarms by 19.61%. This paper presents a proof-of-concept for the feasibility of online transfer-learning on a stream of time-series neurophysiological data that pave the way towards a low-power neuromorphic neuromodulation system.

## 1 Introduction

Harm prevention and mitigation are often far more desirable than dealing with the fallout of deleterious events. Machine forecasting of future events provides an opportunity to integrate preventative systems across a variety of domains. In healthcare, disease prevention is preferable over disease management for both better patient outcomes and for medical resource management, and the same goes for symptoms. In general, the performance of supervised machine learning models are subject to the quantity and quality of training data [1]. This poses a major challenge in healthcare where labeled data is often lacking, and generalization across patients can be difficult to achieve and quantify [2, 3, 4, 5]. In many instances, labeled data is only available at the time of the event, or a brief period of time preceding the event [6, 7]. This significantly limits the flexibility of forecasting models. The relative scarcity of labeled datasets for event prediction and forecasting results in the under-performance of machine learning models for the early detection of many tasks [8, 9, 10].

In this paper, we present a novel and computationally self-sufficient forecasting artificial intelligence (AI) system called ‘AURA’ in the context of physiological signal recording and stimulation, which trains a forecasting network using unlabeled, real-time data that relies on a detection network to provide autonomously generated labels. This is a special case of semi-supervised learning [11, 12], where the Bayes error of the fixed network (detection) is assumed to be less than that of the adjustable network (forecasting), even if the latter were to be considered ‘perfectly’ trained. In AURA, the temporal distance between detection and forecasting tasks allows this assumption to hold. Several principles are utilized in using AURA:

- **Detection outperforms prediction:** In many applications, the performance of a forecasting model degrades as the time between prediction and event (or absence of event) increases. Indicators and bio-markers of events characteristically strengthen as the onset of an event approaches closer in time. The proposed method exploits this fact and uses a detection system to label data which then trains the forecasting model. This is performed with the expectation that incorrectly labeled data by the detection system is unlikely to be correctly predicted by the forecasting system. So emphasis is placed on enabling the predictive system to converge towards the performance of the detector, rather than pushing towards a potentially unrealistic goal of 100% accuracy. Although the asymptotic error of prediction is not shown to converge to that of detection, we show practical examples that demonstrate it works almost as well.
- **New data is more informative than old data:** Online learning can potentially suffer from catastrophic interference (or catastrophic forgetting), where the onset of new training data ‘overrides’ the latent representations of historical data in a neural network [13]. In certain cases, it may be that new data is more representative of present circumstances, so it is preferable to learn from more recent information. More precisely, time-series data is often non-stationary, and the statistical properties of the incoming data are likely to evolve. Online learning could train a system to adapt to changing patient conditions over time.
- **Patient-specific tuning:** While the above point focuses on ‘new’ data in the temporal sense, it also holds true for ‘new’ patients. A model tuned to an individual patient is likely to perform better for that particular individual over a model trained to generalize across a multitude of patients (as is typically the case for most machine learning models in use today). This observation holds true beyond medical diagnostics, for example, geography-specific weather forecasting. Historically, the cost of manually labeling individual patient data to designing patient-specific models has been prohibitively expensive for large-scale deployment. AURA overcomes this using the semi-supervised approach described in Section 2.5.

### 1.1 Novelty and significance

Designing microelectronic circuits and systems for medical implants and electroceuticals is challenging and bounded by several constraints, such as area dimensions, energy consumption, safety, and the need for continuous or very regular data telemetry [35]. While it is not difficult to find medical devices with different capacities of performing on-chip analog or digital signal processing, active on-chip learning use in the neuromodulation or neuromonitoring domain is at its infancy [36].

Fig. 1 demonstrates four general types of loops in medical devices, as applied to neurotechnologies (Fig. 1(a)). In an open-loop system (Fig. 1(b)), decision-making is reviewed by a trained human expert who may perform visits to reprogram the device. This indicates stimulation parameters such as amplitude, frequency, and duty cycle are pre-identified and are indefinitely fixed for the duration of device operation, unless changed manually. This method lacks individualization, may require additional training to equip doctors with the programming skills to re-parameterize the device, exhausts the battery, and results in a very high frequency of unnecessarily applied neurostimulation. The high count of unnecessary stimulation can be considered equivalent to a high number of false positives in a closed-loop system, and hence causes side-effects that push the efficacy of the system into a degree of diminishing returns [37]. An adaptive closed-loop system (Fig. 1(c)), however, can control its stimulation via a feedback loop using extracted bio-markers. Accepting a slightly higher risk of automation in feature extraction by reducing the significance of *expert-in-the-loop* intervention enables significant energy savings, cost benefits and capabilities relative to conventional systems such as deep-brain stimulation (DBS) [9, 14, 15, 16, 17, 18, 19, 20, 21].

**Figure 1.**
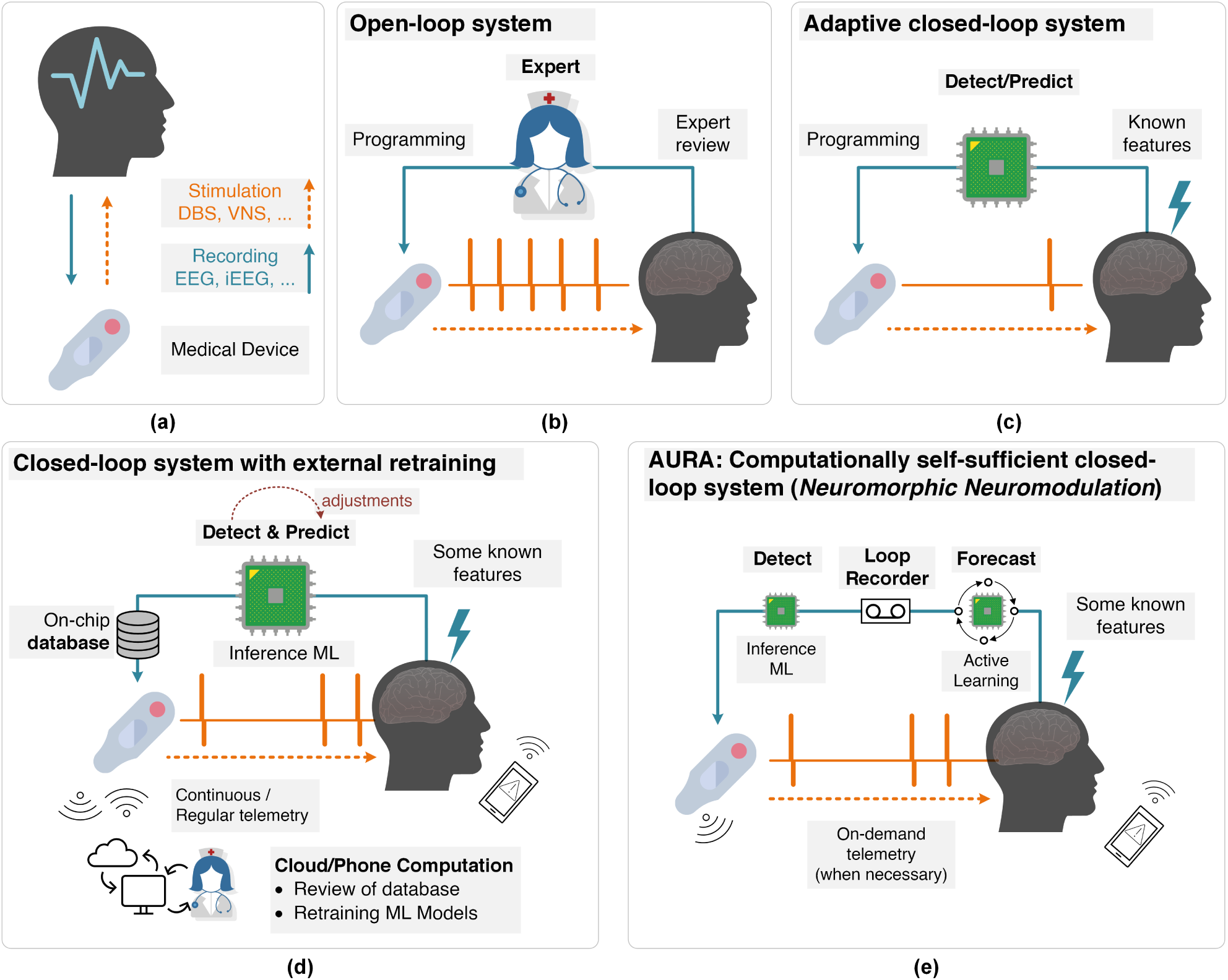
(a) Neuromonitoring (recording) and neuromodulation (stimulation) as part of a medical device and their different system types. While this method in theory can be used with any physiological signal, the most promising are electroencephalogram (EEG), intracranial electroencephalography (iEEG), and local field potentials (LFP), as part of neuromodulation systems such as vagus nerve stimulation (VNS) or deep brain stimulation (DBS). (b) An open-loop system with a human *expert in the loop* that engages occasionally in manual programming and parameter adjustment. (c) An adaptive closed-loop system with an automated bio-marker or feature detection and real-time signal processing that controls simulation. Examples include [9, 14, 15, 16, 17, 18, 19, 20, 21]. (d) An example of a *cloud or smartphone in the loop* system that allows for online adjustments of detection or prediction models, but retraining of the models using the stream of incoming data must be conducted on the cloud. These systems rely on either continuous or regular data telemetry of the data stored in the database. Examples include [22, 23, 24, 25, 26, 27, 28, 29, 30, 31, 32]. (e) AURA operates on the basis of a *neuromorphic neuromodulation system*, where the medical device hosts online training and active learning without any reliance on external computing resources. *Approximate learning* is used to train the active learning model, which pushes the performance of the probabilistic forecasting system towards the non-patient specific inference machine-learning detection model. Once implemented on a low-power neuromorphic chip, system personalization is achieved via its computationally self-sufficient re-training of the forecasting model without the need for data telemetry to the outside world. There are an increasing body of evidence that highly customized low-power neuromorphic systems and models can deliver our vision [33, 34].

Fig. 1(d) illustrates how novel adaptive closed-loop systems are addressing the shortcomings of standard closed-loop systems by incorporating a more complex system architecture with heavy reliance on external computational power to manage control algorithm and feedback signal optimization [20, 38]. These system models that can still be identified as *cloud/expert-in-the-loop* or *phone/patient-in-the-loop*, require continuous or, if pre-identified, regular data telemetry for their machine learning models to be trained on the cloud or smartphone. Many works can be identically or partially identified as offloading resource intensive tasks externally, with reliance on distributed software at the heart of these closed-loop systems [22, 23, 24, 25, 26, 27, 28, 29, 30, 31, 32]. To list a few examples, the role of a processing unit, in innovations such as [32], is limited to the controller of system operation. Another example is outsourcing training, improvement, and optimization of on-chip models to the cloud by regular and continuous transfer of device database [30]. It should be noted that some of the aforementioned methods may offer active adjustments to the system on-the-fly, but none enable on-chip training and re-training, which should be at the core of a successful and highly personalized seizure forecasting system.

Our alternative is ‘AURA’, shown in Fig. 1(e). We believe the combination of an on-chip and actively evolving forecasting model with a *brain loop-recorder* and a dependable detection model, provides a non-trivial, novel, complex and yet, elegant adaptive and personalized system with innovative potential for closed-loop neuromodulation. The alternative we are offering is ambitious with the vision of removing the need for continuous and demanding data telemetry to and from external entities from the loop. The AURA platform still allows for on-demand data telemetry for system diagnostics and performance monitoring.

Personalization in bio-marker discovery and response to therapies continue to demonstrate its profound significance in the field, but it is poorly understood [39, 40]. With respect to seizures, studies in drug-resistant focal epilepsy shows multi-dimensional individualization not only across different patients, but also between seizures in one patient [40, 41]. Personalization is also reported in studies concerning seizure cycles, while more should be done to properly address possibilities for seizure triggers, and accommodate protocols associated with circadian studies [24, 42, 43, 44, 45, 46].

To assess the performance of AURA, we have identified seizure forecasting as an optimal use-case, illustrated in Figure 1. Firstly, the early prediction of a seizure allows for preventative action to take place, such as closed-form feedback via neurostimulation. Secondly, early warning signs manifest in different ways across patients, such as varying brain-wave patterns, and adapting a network to learn the neural signature of a patient could lead to better prediction results. Finally, the largest publicly available labeled scalp electroencephalography (EEG) seizure dataset lacks sufficient information prior to the onset of seizures [6]. The absence of such data means seizure forecasting has been a challenging task for deep learning models, and there is a need to develop techniques that can train and tune models from unlabeled (or indirectly labeled) datasets as well. AURA is perfectly poised to fill this void.

### 1.2 Towards on-chip online-learning for neuromorphic neuromodulation systems

AURA is envisioned as the first step toward designing a patient-specific and highly customized low-power neuromorphic neuromodulation system (nRNS), where sufficient performance for a given patient may justify closed-loop neurostimulation for seizure avoidance. Developing a continuously updated, always-on neuromorphic system that adapts to a patient’s bio-markers requires a low-power, portable learning system that can be ambiently deployed for outpatient care [47, 48]. Deep learning, and particularly training models, is notorious for its large energy consumption which arises due to the huge number of parameters in large-scale models [49, 50, 51, 52, 53], and corresponds to frequent memory accesses and data movement across a chip and between chips. While our prototypical demonstration of AURA provides much promise in the potential of targeted deep learning models in seizure forecasting, extending usage beyond inpatients requires overcoming the energy and latency bottlenecks of on-chip learning [54, 55, 56].

The fields of neuromorphic computing and emerging memory technologies are ushering in new algorithms and architectures that demonstrate how modern deep learning and neuromorphic event-driven systems can be modified to cater for resource-constrained environments. For example, the use of spiking neural networks can reduce the power consumption of equivalent deep learning algorithms by several orders of magnitude [57, 58, 59, 60], and in-memory computing architectures that co-locate processing with network parameter storage can achieve similar performance improvements. Although online/transfer learning and real-time variants of backpropagation algorithms are not often used in practice, AURA as applied to at-risk outpatient tuning motivates a compelling use-case for new generation, online techniques for handling deep learning algorithms, tailored to highly-customized low-power neuromorphic neuromodulation systems [33, 34, 61]. Event-driven neuromorphic computing allows tremendous energy efficiency in both training and inference, but the focus has been primarily on inference engines due to its ubiquity in the IoT (Internet of Things). Recently, significant energy reduction in edge-based training has been demonstrated, where an online transfer learning image processing neuromorphic chip is reported to operate on 23.1 mW (23.6 mW) with 0.8 V supply voltage at 20 MHz, while processing 93k 28 ×28 images/s (100k 28× 28 images/s) during training (inference) [33]. While the simplicity of the network reported here may not be suitable for complex neurophysiological data processing at this point in time^1^, a highly-customized low-power neuromorphic neuromodulation system is a promising avenue to implement and deliver this paper’s vision beyond power-hungry complex deep learning models.

## 2 Background

### 2.1 Seizure Forecasting

It was long thought that epileptic seizures were abrupt events that would materialize without prior warning [63], but the advent of long-term EEG recordings changed this. In the early 1970s, it was shown that seizures could develop over long time scales [64] which pointed to seizure forecasting potentially being within reach. Since then, evidence has amassed to show that seizures are often preceded by detectable changes in brain activity [65, 66], where, for example, a significant increase in blood flow occurs within the epileptic hippocampus prior to temporal lobe epilepsy [67, 68].

An early seizure warning system could improve patient quality of life by triggering pre-emptive administration of therapies, such as anti-epilepsy medication or electrical stimulation [69], which could avert impending seizures and minimize risk of injury. The minimum time interval between an alarm being raised and the occurrence of the seizure while still rendering an intervention to be possible is known as the seizure prediction horizon (SPH).

### 2.2 Detection is easier than prediction

Detecting a seizure at the time of or immediately after onset has had far more success over forecasting seizures in advance [70]. Several machine learning techniques have been used to augment neurologist readings, leading to faster conclusions whilst maintaining specialist-level EEG-reading performance for the identification of seizures as they happen [71, 72, 73, 74]. Unfortunately, seizure aversion is no longer an option when relying on detection mechanisms alone. There are several challenges that face early onset seizure prediction:

- onset patterns vary greatly between patients [75];
- pre-ictal recordings in one patient can be very similar to non-seizure recordings in another patient [76];
- the transition to a pre-ictal state consists of subtle changes that can easily go undetected [77].

The first two challenges highlight the difficulty of developing techniques that generalize across patients. These can be addressed by integrating precision medicine techniques that adapt predictive models tailored to the needs of individual patients.

The third point relates to the challenges of function estimation in temporal data analysis. In deep learning, the goal is to learn a function that maps an input (EEG signals) to an output (whether a seizure will occur after the SPH). The output is treated as a random variable. As the SPH increases, the signal of measurable bio-markers is reduced causing the variance of the output to increase. An intuitive interpretation is that the unpredictable component of the output dominates the predictable component as the time window of forecasting is increased. Reducing the variance of deep learning models can generally be achieved by gathering more data. Unsurprisingly, unlabeled data is far more accessible than labeled data.

### 2.3 The bulk of medical data is unlabeled

Patient-specific models and training with large datasets are somewhat conflicting notions. Individual patients cannot contribute the same scale of data as a whole population of patients. While the availability of long-term EEG recordings has renewed interest in designing patient-specific forecasting algorithms [78, 79, 80], these algorithms still underperform when compared to seizure detection.

The world’s largest public seizure database, the Temple University Hospital (TUH) seizure corpus, contains EEG recordings of over 4,000 patients, and is commonly used for testing and validating the performance of seizure detection systems that achieve close to expert clinician performance [6, 71]. The recordings commence between 5 to 35 minutes pre-ictally which limits the length of the SPH. While there is a plethora of unlabeled data, the lack of formally trained EEG readers available to provide precise temporal annotations, with confirmatory secondary readings, means this huge source of data cannot be directly used in supervised learning methods. The bulk of medical data remains unlabeled and underutilized.

This challenge can be addressed by relying on high performance seizure detection models to annotate this data for us. Weak supervision has previously been applied by obtaining inaccurate labels from a mix of experts and novices for real-time seizure detection [81]. Our approach can be distinguished as we wholly do away with manual annotations, and instead use a detection model shown to perform similarly to neurologists [71]. These machine-generated detection labels are then used as targets for the prediction model. Some of the detected seizures may be misclassified (and thus, inaccurate labels for prediction), which raises concerns that noisy labels could mislead the prediction system [82].

Fortunately, in the real world, the fact that temporal data is inherently correlated is extremely useful. A seizure that is misclassified at its onset (real-time detection) is unlikely to be successfully predicted pre-ictally (forecasting), as the unpredictability of seizures generally increases with a longer SPH. Therefore, such errors are treated as inevitable; the larger amount of correctly classified data is instead used to offset potential performance degradation from noisy labels. Training a prediction system using approximate labels derived from a detection system makes better use of temporal correlations in the real world [82, 83, 84].

### 2.4 Learning patient-specific patterns

Seizures may be regarded to follow patient-specific cyclic patterns. This has been long observed since 1939, when Griffiths and Fox observed that some patients experience seizures at certain times of the day, while others followed monthly cycles [85]. Confounding variables also contribute to the variance between patients, including medication, stress, circadian effects, and hormonal effects, amongst others [86, 87, 88, 89]. Circadian (days) and multidien (multi-days) seizure cycles have also recently been studied with patient self-reported diaries and retrospectively on some long-term intracranial EEG data, which shows peaks in seizure cycles as long as 30 days apart [43, 90, 91, 92]. Whether the observed cycles are valid and whether they can be linked to triggers such as missed medication, mental and emotional states, the menstrual cycle, and the duration and severity of seizures require objective and prospective studies [93]. It is known that mammalian physiology and behavior is widely influenced by light and other environmental factors, which means circadian or multidien studies require well-developed protocols before the study is conducted [94].

This poses a challenge in developing models that generalize across populations. The present approach to designing personalized forecasting models relies on individualized labeled data, which demands precise temporal annotations for each future time window during training. While this may be feasible for small-scale datasets, it is not a long-term tenable solution for challenging tasks, such as seizure forecasting, or for patient-specific tuning with a large population of patients.

### 2.5 AURA

This work proposes AURA, an adaptive, unlabeled, real-time and approximate approach to online learning, and its performance is demonstrated on patient-specific seizure forecasting. An overview of AURA as applied to seizure forecasting using several datasets procured across three different continents is illustrated in Figure 2 (described in further detail in Methods). AURA consists of a real-time detection network and a forecasting network. The detection network classifies the onset of seizures in real-time with acceptable accuracy, and the forecasting network uses the output of the detection network as labels. This allows training to take place with the plethora of unlabeled data that is available, and pre-trained forecasting networks can be retrained to adapt to individual patients in order to learn patient-specific pre-ictal signatures.

**Figure 2.**
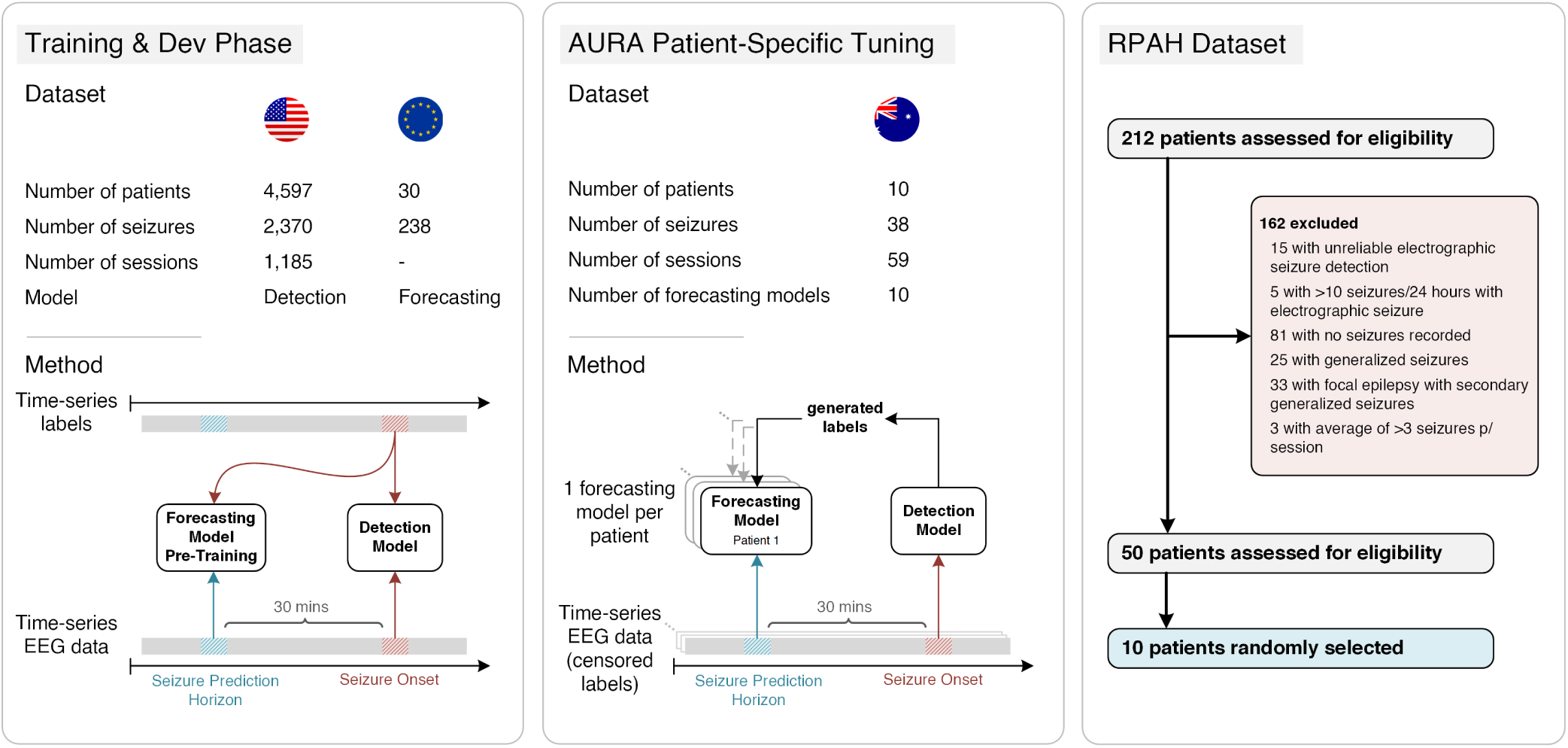
AURA Methodology as applied to seizure prediction. The detection model is trained using a U.S. dataset, and the prediction model is pre-trained with an EU dataset. The AURA patient-specific tuning phase assigns a replica of the pre-trained prediction model to each patient from the AU dataset, and all ground-truth labels are censored. The global detection model generates labels in real-time which are used to train the 10 patient-specific prediction models across 59 sessions of EEG recordings. Each patient has a personalized forecasting model that has been tuned to their pre-ictal signatures. To provide a measure of forecasting performance, the labels are declassified and compared to the predictions generated during the AURA patient-specific tuning process.

This approach is a narrow form of semi-supervised learning, where a pre-trained detection model achieves acceptable performance on a modestly sized dataset, while the forecasting network is updated using initially unlabeled data. Although inaccurately labeled data may deceive the forecasting network, AURA is implemented with the inductive prior that the forecasting network is unlikely to predict what fails detection. The irreducible error of the detection model must therefore be less than that of the prediction model, which is a reasonable assumption for temporally correlated prediction and detection. Performance degradation is compensated for by expanding the pool of usable data by the forecasting network for training.

We perform a pseudo-prospective study using AURA across 10 patients. The detection network is trained on patients from a U.S.-based hospital, which generates labels during the AURA online learning process on a sample of patients from the Royal Prince Alfred Hospital (RPAH) in Sydney, Australia. The AURA-trained forecasting networks show an average improvement in relative sensitivity by 14.30%, and a reduction in false alarms by 19.61%. A high-level overview of the AURA training process is depicted in Figure 2.

## 3 Datasets

The seizure detection model is trained on the Temple University Hospital (TUH) seizure corpus [95] from the U.S., while the prediction model is pre-trained using the European (EU) EPILEPSIAE dataset [96]. The AURA self-learning process is applied to the Australian test set from the RPAH where all human-annotated labels have been censored [71], and each patient starts with the same pre-trained prediction model that adapts over the course of their multiple monitoring sessions. Upon completion of all sessions, the sequence of predictions generated by the forecasting network is compared to the uncensored ground truth to provide a performance measure of sensitivity and the number of false alarms. It is important to note that each prediction for a given time window takes place 30 minutes prior to the detection network generating a label that corresponds to the same window. This ensures our metric for performance using any given sample is reported before the forecasting network is updated on the basis of the same sample. This reduces the risk of future time leakage into our measure of performance, and emulates the operation of AURA in practice.

### 3.1 TUH dataset

The TUH dataset [95] is the world’s largest open EEG database for seizure research. It includes 592 patients in the training dataset and 50 patients in the test set. Due to the lengthy sessions, much of the recordings that do not include seizure onset have been removed. The temporal discontinuity of the data and the lack of sufficient pre-ictal content means the TUH dataset is not suitable for use with AURA, and would not be representative of real-world usage. However, it is an ideal dataset for training the seizure detection model, despite the seizure and background information imbalance. The details are shown in Table 1, in which the total seizure and background duration in the Train/Dev datasets are 46.7 h and 752.3 h, respectively. Public access to TUH dataset is possible via online registration and application for access.

**Table 1.** Summary of TUH dataset

| Attribute | Train/Dev (80/20) | Test |
| --- | --- | --- |
| Files | 4597 | 1013 |
| Sessions | 1185 | 238 |
| Patients | 592 | 50 |
| Files with seizures | 867 | 280 |
| Sessions with seizures | 343 | 104 |
| Patients with seizures | 202 | 40 |
| Number of seizures | 2370 | 673 |
| Background duration (hours) | 705.6 | 154.1 |
| Seizure duration (hours) | 46.7 | 16.2 |
| Total duration (hours) | 752.3 | 170.3 |

\*Training the detection model follows the same procedure as in [71]. The TUH seizure corpus provides dedicated validation and test sets, although the labels from the test set are unreleased. Therefore, the default validation set was instead used as a held out test set for performance benchmarking of the detection model, while the train set was randomly split with a ratio of 80-20 to create our own validation subset.

### 3.2 EPILEPSIAE dataset

The EPILEPSIAE dataset is the largest continuous EEG database in Europe which contains a total of 275 patients [96], among which, scalp-EEG recordings taken from 30 patients are made publicly available. Although this number is much less than the TUH dataset, the recordings of all patients are significantly longer in duration, ranging between [92.9, 266.4] hours. A summary of patient details are provided in Table 2 with a total of 238 seizures across a 4604 h recording duration. This dataset is used to pre-train the seizure prediction model. Public access to EPILEPSIAE dataset is possible and requires payment, registration and application for access.

**Table 2.** Summary of EPILEPSIAE scalp-EEG dataset.

| Patient | Gender | Age Range | SN | Seizure-Foci | RD (h) | Mean SD (s) | Range SD (s) |
| --- | --- | --- | --- | --- | --- | --- | --- |
| 1 | M | 36 – 40 | 11 | Central & Parietal | 164.7 | 78.3 | [54.5, 162.0] |
| 2 | F | 46 – 50 | 8 | Temporal | 177.4 | 52.9 | [4.3, 71.3] |
| 3 | M | 40 – 45 | 8 | Temporal | 143.3 | 58.6 | [34.3, 102.8] |
| 4 | F | 66 – 70 | 5 | Temporal | 167.8 | 121.0 | [84.3, 166.1] |
| 5 | F | 50 – 55 | 8 | Frontal & Temporal | 266.4 | 92.3 | [11.3, 379.1] |
| 6 | M | 60 – 65 | 8 | Temporal | 135.4 | 50.2 | [0.0, 105.1] |
| 7 | M | 36 – 40 | 5 | Temporal | 118.1 | 46.7 | [29.3, 79.6] |
| 8 | M | 26 – 30 | 22 | Frontal | 115.6 | 20.1 | [0.9, 96.3] |
| 9 | M | 46 – 50 | 6 | Temporal | 94.0 | 71.4 | [65.9, 83.8] |
| 10 | M | 40 – 45 | 11 | Frontal & Temporal | 138.0 | 39.9 | [0.0, 68.1] |
| 11 | M | 46 – 50 | 14 | Central & Temporal | 138.1 | 63.6 | [19.4, 100.4] |
| 12 | M | 26 – 30 | 9 | Temporal | 159.7 | 41.4 | [31.2, 60.8] |
| 13 | M | 46 – 50 | 8 | Frontal & Temporal | 158.1 | 91.0 | [55.0, 125.5] |
| 14 | F | 60 – 65 | 6 | Temporal | 162.2 | 124.0 | [83.0, 174.1] |
| 15 | F | 40 – 45 | 5 | Temporal | 118.7 | 64.9 | [7.6, 144.0] |
| 16 | F | 10 – 15 | 6 | Temporal | 92.9 | 56.1 | [2.5, 99.0] |
| 17 | F | 16 – 20 | 9 | Temporal | 159.1 | 55.8 | [33.3, 76.5] |
| 18 | M | 46 – 50 | 7 | Temporal | 178.2 | 41.7 | [22.7, 69.4] |
| 19 | M | 30 – 35 | 22 | Temporal | 161.1 | 65.2 | [43.1, 96.5] |
| 20 | M | 46 – 50 | 7 | Temporal | 164.6 | 59.5 | [19.1, 119.8] |
| 21 | F | 30 – 35 | 8 | Occipital | 159.4 | 51.9 | [9.3, 118.9] |
| 22 | M | 36 – 40 | 7 | Parietal | 137.9 | 95.7 | [55.5, 145.1] |
| 23 | M | 50 – 55 | 9 | Temporal | 237.5 | 56.1 | [18.8, 88.0] |
| 24 | F | 50 – 55 | 10 | Temporal | 94.4 | 72.9 | [49.6, 122.8] |
| 25 | M | 40 – 45 | 8 | Central | 159.7 | 329.3 | [73.8, 909.2] |
| 26 | M | 10 – 15 | 9 | Temporal | 159.0 | 62.7 | [38.0, 104.5] |
| 27 | M | 56 – 60 | 9 | Temporal | 159.5 | 95.3 | [43.9, 331.2] |
| 28 | F | 30 – 35 | 9 | Temporal & Parietal | 162.3 | 86.2 | [59.4, 216.9] |
| 29 | M | 50 – 55 | 10 | Temporal | 161.1 | 180.7 | [62.1, 270.0] |
| 30 | F | 16 – 20 | 12 | Temporal | 159.8 | 75.2 | [10.0, 151.9] |
| Total | — | — | 238 | — | 4604 | 75.9 | [0.0, 909.2] |

M: Male, F: Female, SN: Number of seizures, RD: Recording duration, Mean SD: Mean of seizure duration, Range SD: Seizure duration range

### 3.3 RPAH dataset

Our selection procedure is detailed in Fig. 2. There are a total of 212 adult patients with surface EEG recordings at the RPAH (Sydney, Australia), where 192 patients have reliable EEG recordings (excluding 15 patients that are lacking full electrode information, and other 5 patients that experience more than ten seizures/24 hours). Among the remaining potential subjects, 111 patients have seizures recorded. The pool of patients is narrowed to those with focal epilepsy who: i) do not experience generalized or secondary generalized seizures, as the initial source of each seizure varies, ii) experience at most three seizures per day on average to provide sufficient inter-ictal training time (Figure 3a), and iii) have at minimum three sessions (~ three days) recorded. 50 patients are assessed for eligibility; among these patients, 10 patients are used as subject-under-test for real-time AURA training across a total duration of 949.9 hours. Detailed information for each patient is provided in Table 3. Each patient starts with an identical pre-trained forecasting model, which is adapted during the AURA process.

**Table 3.** Summary of RPAH selected dataset

| Patient | Gender | Age Range | Seizure Num. | Session Num. | RD (h) | Mean SD (s) | Range SD (s) | Seizure Type | Seizure Foci | Secondary Generalized |
| --- | --- | --- | --- | --- | --- | --- | --- | --- | --- | --- |
| 1 | F | 20 – 25 | 3 | 7 | 116.3 | 50.5 | [47.3, 53.6] | Focal | Left Occipital | N |
| 2 | F | 50 – 55 | 4 | 6 | 95.1 | 70.4 | [29.6, 94.0] | Focal | Right Frontal | N |
| 3 | M | 40 – 45 | 1 | 4 | 70.6 | 118.3 | [118.3, 118.3] | Focal | Left Temporal | N |
| 4 | F | 46 – 50 | 3 | 7 | 158.8 | 76.7 | [65.8, 92.8] | Focal | Left Parieto-Occipital | N |
| 5 | F | 30 – 35 | 7 | 5 | 53.7 | 61.1 | [36.2, 91.3] | Focal | Left Temporal | N |
| 6 | M | 46 – 50 | 6 | 8 | 94.3 | 48.0 | [36.5, 56.4] | Focal | Right Temporal | N |
| 7 | M | 66 – 70 | 7 | 5 | 55.9 | 55.6 | [46.7, 67.7] | Focal | Left Temporal | N |
| 8 | M | 46 – 50 | 2 | 4 | 46.9 | 64.7 | [58.2, 71.2] | Focal | Right Temporal | N |
| 9 | M | 60 – 65 | 2 | 8 | 162.6 | 62.1 | [59.4, 64.8] | Focal | Right Temporal | N |
| 10 | F | 30 – 35 | 3 | 5 | 95.2 | 219.3 | [138.0, 363.7] | Focal | Right Temporal | N |
| Total | — | — | 38 | 59 | 949.4 | 73.6 | [29.6, 363.7] | — | — | — |

M: Male, F: Female, SN: Number of seizures, RD: Recording duration, Mean SD: Mean of seizure duration, Range SD: Range of seizure duration, Secondary Generalized: Secondary generalized seizures refer to those that initiate focally and end with bilateral motor activity. They are often clinically classified as focal seizures.

**Figure 3.**
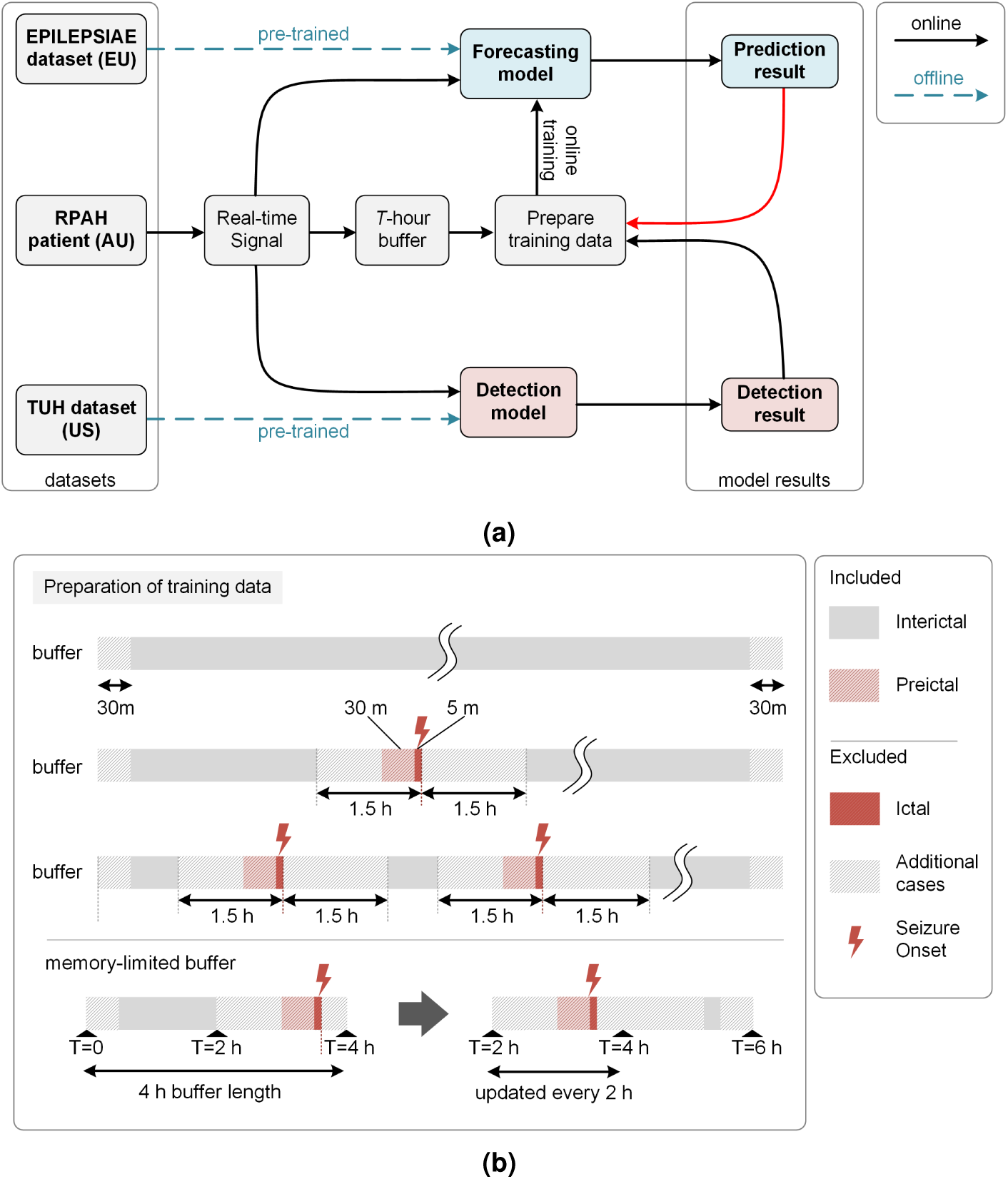
Online learning procedure. (a) The prediction and detection models are initially pre-trained offline. The EEG recording from a given patient from the RPAH dataset is streamed as a real-time signal. It is used to generate a real-time detection (detection result) and a 30-minute forecast (prediction result). The detection result i) determines whether the sample is used to train the prediction model, and ii) is used as the target label for the prediction model outcome from 30-minutes prior. The red arrow between ‘prediction results →prepare training data’ indicates that prediction results can be used to govern the training of the prediction model; e.g., if the prediction results are aligned with the detection results, there is no need to update the prediction model. (b) Training data preparation. If the detection result indicates an inter-ictal or pre-ictal (within 30 minutes of onset) reading, online training of the prediction model is enabled. If the reading is ictal (within 5 minutes of the seizure onset), post-ictal (within 1.5 hours after seizure onset), online training is disabled as the signal is considered ‘contaminated’. Training is disabled during the first 30 minutes of the buffer due to the absence of corresponding forecast labels, and also during the final 30 minutes of the buffer as the prediction results fall outside of the range of the buffer. To ensure forecasted seizures in the final 30 minutes are still accounted for, incoming data sequentially replaces the oldest data in the buffer, emulating the function of a loop recorder. The minimum usable buffer size must account for the 30-minute pre-ictal duration, the 1.5-hour gap between ictal and inter-ictal period, 30-minutes at either end of the buffer, and the seizure duration itself. Our experiments use a 4 hour buffer length.

This study on the RPAH clinical data is approved by the local Research Ethics Committee. Ethics approval number *X* 19-0323-2019/STE16040 on *Validating epileptic seizure detection, prediction and classification algorithms* approved on 19 September 2019 by the NSW Local Health District (LHD) for implementation at the Comprehensive Epilepsy Services, Department of Neurology, The Royal Prince Alfred Hospital (RPAH). The RPAH data is not openly available to the public.

## 4 Methods

### 4.1 Seizure prediction using AURA online learning

The online learning process using real-time streamed EEG data is shown in Figure 3. The surface EEG signal of a given patient is read into the buffer in Figure 3(a) once per second, where it is stored until it reaches *T* hours. In our implementation, our buffer was limited to *T* = 4 hours. Once the total duration of the recording exceeds *T*, new data is loaded into the buffer while earlier data is sequentially cleared which ensures positive forecasts in the final 30 mins of the buffer are accounted for. Concurrently, the two models generate detection and prediction outputs for batches of 12 s and 30 s signals, respectively. Once the buffer reaches *T* hours, the training data for the prediction model is prepared based on the detection result (weak label), as illustrated in Figure 3(b). EEG signals associated with inter-ictal and pre-ictal (up to 30 mins) data is included in the online training process. Ictal and post-ictal (up to 1.5 hours) data is excluded from the online learning process, as generating forecasts at seizure onset would be based on contaminated signals. The first 30 mins of EEG recordings in the buffer is also excluded as there is no guarantee the patient is not in a post-ictal state, and also due to the absence of forecasted labels in the first 30 mins. The final 30 mins is excluded as there is no knowledge of the state of the patient, e.g., a seizure may occur right after the buffer. This ‘right-censored’ recording is accounted for in subsequent steps once the buffer is updated over time, and the data becomes uncensored by shifting along the buffer’s storage. Once the system has flagged a data sample for inclusion in the online training process, the weak label from the detection model is compared to the prediction result using a negative log-likelihood loss function, followed by a gradient calculation step via the backpropagation algorithm.

#### 4.1.1 Real-time signal pre-processing

Once the patient’s real-time signal accumulates to a 12 second window, independent component analysis (ICA) [97] and short-time Fourier Transform (STFT) are applied to the EEG signal before being passed to the pre-trained seizure detection model. ICA is applied to decompose the signal into several statistically independent components. The electro-oculography (EOG) channel records eye movement information, and is physically proximate to the ‘FP1’ and ‘FP2’ EEG channels. Independent components of noise-prone EEG channels above a defined Pearson correlation threshold with the EOG channel are removed. STFT is then applied to the clean EEG waveform with a 250 sample window (1 second) length and 50% overlap. The DC component of the transform is also removed as it is known to have no relation to seizure occurrences. The same technique is used on the real-time stream of EEG data for the prediction model, but using 30 second windows instead.

#### 4.1.2 Seizure detection and prediction pre-trained models

All pre-training takes place offline, where the seizure detection model is trained using the TUH dataset (see Section 3.1) and the prediction model using the EPILEPSIAE dataset (see Section 3.2). The detection and prediction models both consist of convolutional long short-term memory (ConvLSTM) modules [98] combined with a pair of fully-connected layers, based on our previous work on seizure detection [71]. A summary of the architecture is provided in Table 4.

**Table 4.** Network Architecture

| Layer | Type | Parameters |
| --- | --- | --- |
| 1 | ConvLSTM | $k = (n \times 2 \times 3), f = 16, s = 1$ |
| 2 | ConvLSTM | $k = (16 \times 1 \times 3), f = 32, s = (1 \times 2)$ |
| 3 | ConvLSTM | $k = (32 \times 1 \times 3), f = 64, s = (1 \times 2)$ |
| 4 | FC | $m = 256$, sigmoid |
| 5 | FC | $m = 2$, sigmoid |

$k$ : kernel dimensions, $n$ : input channel dimensions, $f$ : number of filters, $s$ : stride, FC: fully-connected layer, $m$ : number of neurons

#### 4.1.3 Buffering

In our system, the buffer has a size of four hours of loop EEG recording. Specifically, every two hours, new EEG signals are added to the buffer and old signals are flushed in a first-in-first-out fashion. The buffer also contains timestamps of the EEG signals, which are used to prepare the training data. Note that the size of the buffer (four hours in this work) and the frequency of updating the buffer (two hours in this work) are subject to the available computation and memory resources. In other words, the online training process that uses the buffered data must finish before the next buffer update. This process is depicted in the illustration of the memory-limited buffer in Figure 3b.

#### 4.1.4 Preparation of training data

The forecasting model’s online training data preparation process commences once the buffer reaches *T*− hours (*T* = 4 for our experiments). The feedforward detection model labels all suspicious instances of seizure onset. There are three possible cases that may arise in the 4-hour buffer. **Case 1**: if no seizure is labeled in the T-hour buffer duration, the entire duration of the buffer will be labeled inter-ictal (negative sample), other than the first and final 30 minutes. **Case 2**: if only one seizure is labeled in the *T*− hour buffer, the 30 minutes preceding the seizure is labeled pre-ictal (positive sample), and any information that falls 1.5 hours away from the seizure onset within the buffer is labeled inter-ictal (negative sample). **Case 3**: if more than one seizure is marked during the *T*− hour buffer, as before, a positive (pre-ictal) label is assigned 30 minutes preceding the seizure, and a negative (inter-ictal) label is assigned at least 1.5 hours away from all seizures. Once the entire buffer consists of labeled EEG information, only those with inter-ictal and pre-ictal labels are further pre-processed using 30 second time windows and STFT as described in 4.1.1.

#### 4.1.5 Online training

During the real-time prediction phase, the prediction model is continuously updated based on the results of the detection labels. An adaptive batch size (varied from 5 to 32) is used, with the exact size based on the number of training samples, noting that even for a fixed buffer size, the criterion in Section 4.1.4 alters the number of samples flagged for training based on the occurrence and timing of seizure alarms. The Adam optimizer is used with a learning rate of 5 × 10^−8^. The purpose of using a small learning rate and batch size is to avoid overfitting caused by the small amount of training data within the *T* −hour buffer.

#### 4.1.6 Post processing

During the online training process, raw prediction results are further post processed by calculating the moving average value during a given period, as shown on the Figure 4. In our experiments, we use a window of 30 minutes. A patient-specific threshold is applied, where if exceeded, an alarm is raised, predicting the patient will experience a seizure within the next hour.

**Figure 4.**
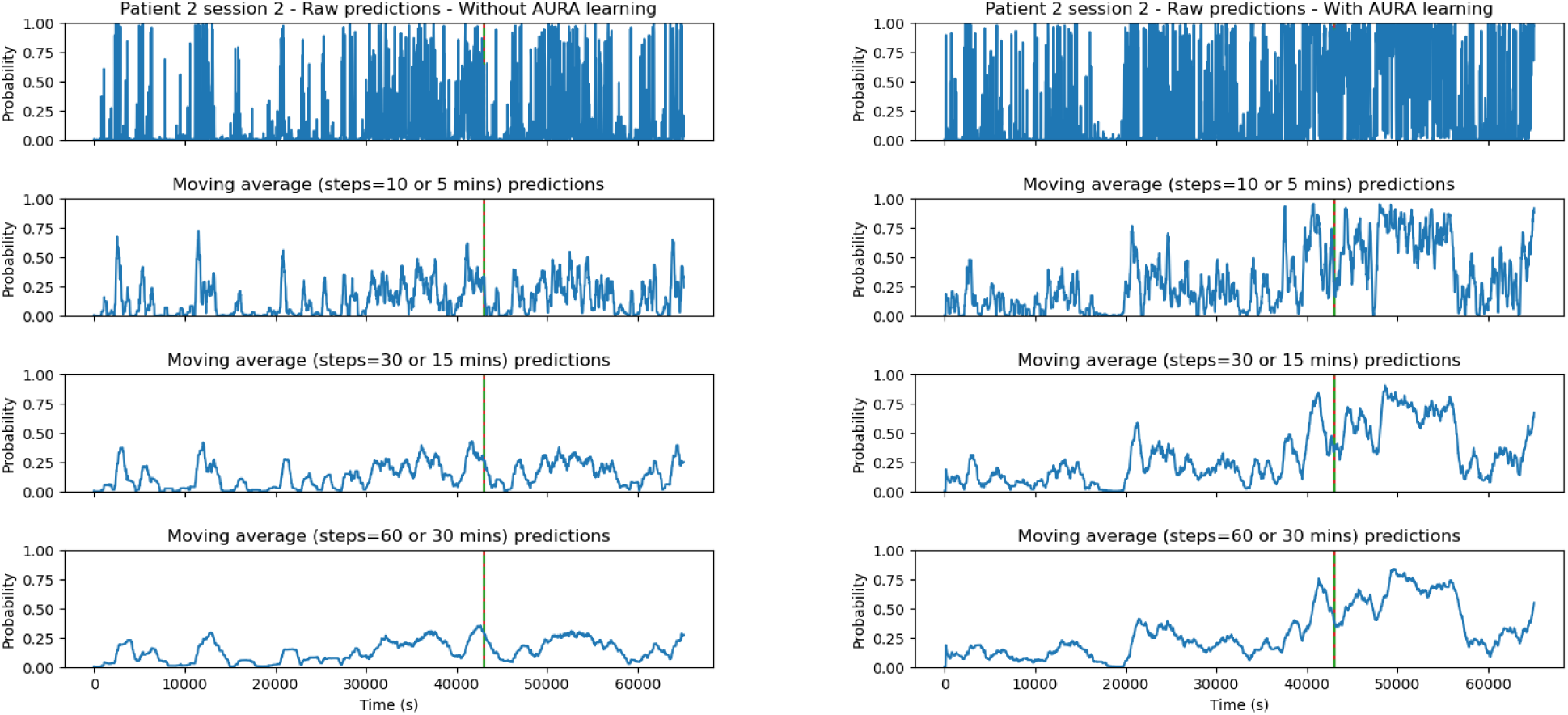
Real-time prediction comparison of a sample patient session without AURA learning (left figure) and with AURA learning (right figure). The dashed vertical red line represents the ground truth seizure onset time, and the solid green line is the weak label generated by the seizure detection model. In the above example, these two lines overlap which means the detection model correctly classified the actual seizure onset. The peak that precedes the seizure event corresponds to the prediction system forecasting a high probability that a seizure will occur within the next hour. The peak in AURA learning is far more distinguishable than that without AURA learning. The sample is taken from the second session of EEG recordings, resulting in different probabilities at *t* = 0 *s* as the model has undergone one session of AURA training.

#### 4.1.7 Performance metrics

The sensitivity and number of false alarms per 24 hours are used to evaluate the performance of AURA in real-time seizure prediction for clinical usage. When the prediction results are greater than a specific patient-based threshold, one alarm is raised. The thresholds are chosen for each patient to achieve a balance between sensitivity and false alarm. An alarm is considered correct if it is raised within one hour of the actual seizure onset. Where multiple incorrect alarms occur within the same hour, they are collectively regarded as one false alarm. Note that during the training process, we define pre-ictal window of 30 minutes as it has empirically proven to be effective in training a seizure prediction system [99]. This temporal discrepancy arises because the timing of pre-ictal bio-marker onset remains an open question.The sensitivity of the prediction network is calculated by finding the number of correctly predicted seizures over the total number of seizures. The total number of false alarms over all recording sessions is normalized to calculate the number of false alarms per 24 hours. The time-in-warning is also calculated to show the amount of time each patient is on high alert, and is calculated by dividing the total time spent in a warning state following an alarm (TP+FP) by the total recording time.

The prediction performance score is also calculated based on the sensitivity and false alarm, inspired by the score formula proposed in the Neureka 2020 Epilepsy Challenge [100]:

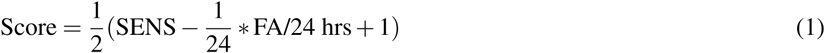

As the predictor alarm is considered to last for an hour, the upper bound on false alarms per 24 hours is 24. The coefficients of the score formula are selected to fit the score within a range of 0 to 1. The Wilcoxon signed-rank statistic is used to evaluate the performance of the AURA system compared with the baseline. To account for both sensitivity and the number of false alarms, the *p*-value is calculated based on the patients’ prediction performance score to indicate the significance of performance improvement. The statistical significance threshold was set to 0.01.

## 5 Results

The following pseudo-prospective tests are applied to the RPAH patients:

- **Detection**: The detection model is pre-trained on the TUH dataset and is pseudo-prospectively inference-only real-time tested on the RPAH dataset.
- **Without AURA**: The prediction model is pre-trained on the EPILEPSIAE dataset and is pseudo-prospectively inference-only real-time tested on the RPAH dataset.
- **With AURA**: The prediction model is pre-trained on the EPILEPSIAE dataset and is trained online using the AURA system, using labels that are simultaneously generated by the pre-trained seizure detection model (teacher model), and pseudo-prospectively inference-only real-time tested on the RPAH dataset.

The recordings of each patient from the RPAH dataset are pseudo-prospectively tested on the prediction model with AURA learning, by streaming their scalp-EEG readings into our system in real-time. Additionally, the same data is used on the pre-trained prediction model (on the EPILEPSIAE dataset) without AURA learning to identify the effectiveness of the proposed framework. As shown in Table. 5, all ten patients show varying degrees of improvement. Patients 1, 2, and 5 have significant improvement, as their sensitivity increases by 33.34%, 25.00%, and 14.28%, respectively, while the number of FA/24hrs decrease by 0.62, 2.01, 0.90, respectively. The result for patient 3 is quite promising as the sensitivity is maintained at 100.00% with approximately 2 FA/24hrs with AURA learning. The probability of the null hypothesis, that AURA does not improve the predictive performance score of the forecasting model without AURA, was calculated to be *p* = 0.003, which is smaller than 0.01. The average result across patients without AURA is 55.26% sensitivity, 8.21 FA/24hrs, and across patients with AURA is 63.16% sensitivity, and 6.60 FA/24hrs. These results are quite promising considering these are non-patient-specific continental generalized tests without human labeling. Other patients 4,6,7,8,9,10 show varying levels of FA/24hrs, but upon AURA training, there is a consistent decrease of both FA/24hrs and time warning, for a constant sensitivity.

Fig. 4 shows a sample taken from the second session of patient 2 experiencing focal epilepsy. The dashed vertical red line shows the ground truth seizure onset time, and the solid green line represents the weak label generated by the seizure detection model. In this case, the detection model correctly labels the actual seizure onset such that they are almost perfectly overlaid. The first row shows the raw prediction results, and the subsequent rows show a moving average of 5, 15, 30 minutes results, respectively. From the graph, we can see that without AURA, the inference-only test shows a very low probability of seizure occurrence, while with the AURA training process, the model becomes more sensitive to seizures by showing a higher probability of an alarm being raised prior to seizure onset. This is achieved concurrently with reducing the number of FA/24hrs for each patient.

## 6 Discussion

In this study, we present the AURA online learning system that uses patient-specific semi-supervision with a pair of temporally correlated tasks, and demonstrate that it consistently improves (or maintains) patient-specific seizure prediction results in terms of both sensitivity and the number of false alarms. Based on the average of ten patients’ results, the real-time seizure detection model outperforms the prediction model, which reaches an average of 73.68% sensitivity, 4.17 FA/24hrs, and only 0.50% time warning. Averaging the sensitivity and FA/24hrs across all patients shows the prediction model achieves 63.16% sensitivity, 6.60 FA/24hrs with AURA learning, and 55.26% sensitivity, 8.21 FA/24hrs without AURA learning. With the AURA system learning, the seizure prediction model moves closer to the performance of the detection model. We can see that the seizure prediction result improves across all ten patients with only several sessions per patient, with an absolute average improvement of sensitivity by 14.30%, and an average reduction of 1.61 FA/24hrs. The duration a patient is in time warning also decreased by 16.67%, which can reduce alarm fatigue in clinical settings.

### 6.1 Adapting to patient-specific biomarkers

In 2001, a five-patient clinical study was conducted that suggested epileptic seizures commence after a cascade of electrophysiological events which occur far earlier than clinical onset [101]. Our recent work [102] postulates that slowing inter-ictal activities are a potential biomarker for epileptic seizure prediction. The gradual improvement of results using AURA, to some extent, supports the hypothesis that patient-specific early warning signs are regularly raised before the seizure onset.

The three patients with the largest margin of improvement (1, 2 and 5) interestingly have focal epilepsy with different specific seizure foci: left occipital, right frontal and left temporal, respectively. Additional details are provided in Table 3. Thus, for patient-specific online training, if a biomarker exists and the detection model can continuously and correctly generate labels of seizure onsets, the forecasting model can theoretically learn to identify seizure prediction biomarkers for that specific patient.

### 6.2 Dataset Generalization

AURA is demonstrated on an out-of-distribution dataset to what the detection and prediction models are pre-trained on, and is representative of different populations and recording practices. There are two instances where the system is required to generalize in our experimental system. Firstly, the detection model is trained on the U.S.-based TUH dataset but must generate labels for the Australian RPAH dataset. Secondly, the prediction model is initialized with pre-trained parameters from the European dataset, but performance is reported on the Australian patients. This means that when the prediction model reports (or misses) the *first* pre-ictal stage in the first session for any given patient, it will take another 30 minutes before the model is exposed to its first sample of seizure onset data from the RPAH dataset. Therefore, performance is reported 30 minutes prior to updating the model via backpropagation using that particular prediction in the calculation of the loss. This reduces the risk of future-time leakage.

The performance of the detection model for each patient is also reported in Table 5, where the average performance of detection is better than that of the prediction system with AURA learning. While this result is unsurprising, i.e., AURA is implemented on the basis that the upstream task of detection is easier than the downstream task of prediction, it is interesting to see AURA leads to better forecasting for patients 1 and 3 in both sensitivity and FA/24hrs than detection. The detection model evidently does not impose an upper-bound on performance, but this counterintuitive result may be attributed to the detection network struggling to generalize. For example, if pre-training of the detection network were to take place on the same RPAH cohort, then the irreducible error of the detection system would likely be less than that of the prediction network, even with AURA learning. This is consistent across all ten patients, who experience seizure prediction performance that moves closer to that of detection. The patients show varying levels of improvement, and more patient tests and experiments from a wider variety of seizures types must be implemented to instill higher confidence of the proposed system.

**Table 5.**
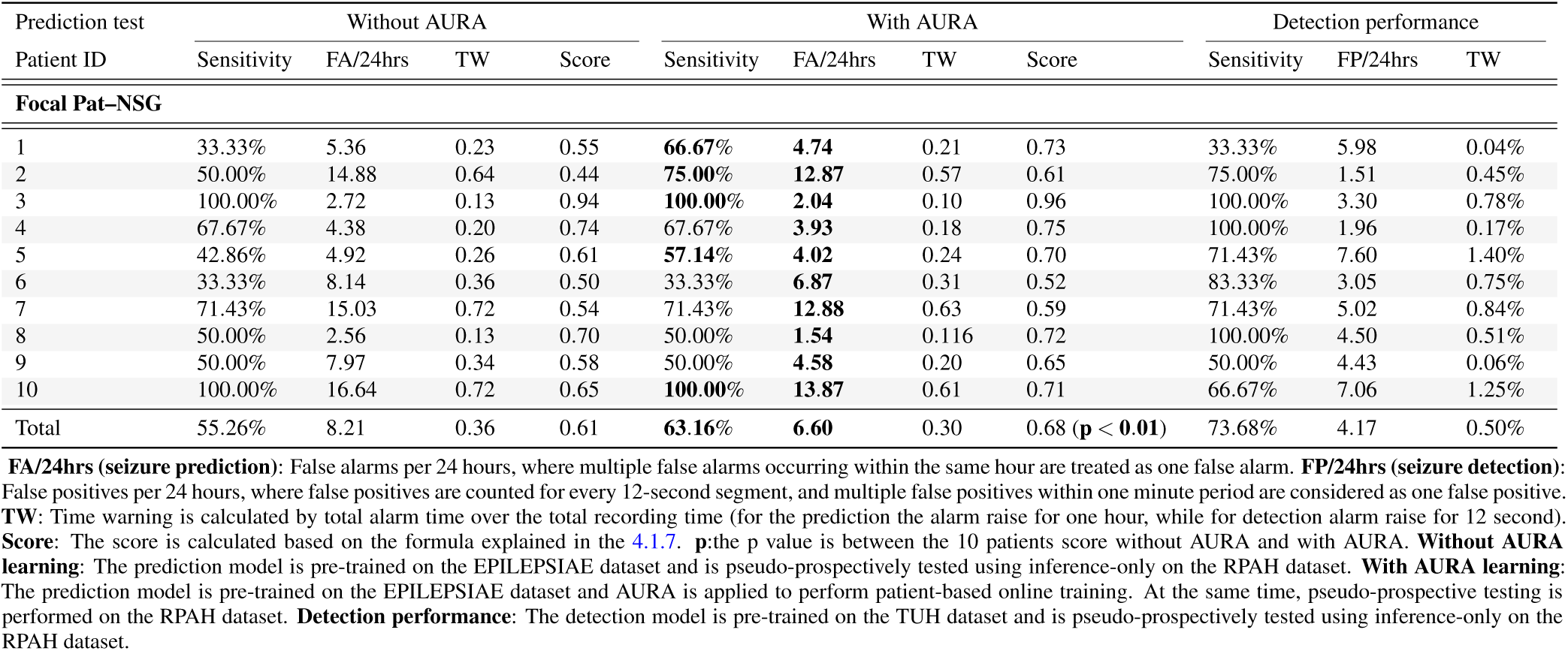
Continental generalization pseudo-prospective results comparison

| Prediction test | Without AURA |  |  |  | With AURA |  |  |  | Detection performance |  |  |
| --- | --- | --- | --- | --- | --- | --- | --- | --- | --- | --- | --- |
| Patient ID | Sensitivity | FA/24hrs | TW | Score | Sensitivity | FA/24hrs | TW | Score | Sensitivity | FP/24hrs | TW |
| **Focal Pat-NSG** |  |  |  |  |  |  |  |  |  |  |  |
| 1 | 33.33% | 5.36 | 0.23 | 0.55 | **66.67%** | **4.74** | 0.21 | 0.73 | 33.33% | 5.98 | 0.04% |
| 2 | 50.00% | 14.88 | 0.64 | 0.44 | **75.00%** | **12.87** | 0.57 | 0.61 | 75.00% | 1.51 | 0.45% |
| 3 | 100.00% | 2.72 | 0.13 | 0.94 | **100.00%** | **2.04** | 0.10 | 0.96 | 100.00% | 3.30 | 0.78% |
| 4 | 67.67% | 4.38 | 0.20 | 0.74 | 67.67% | **3.93** | 0.18 | 0.75 | 100.00% | 1.96 | 0.17% |
| 5 | 42.86% | 4.92 | 0.26 | 0.61 | **57.14%** | **4.02** | 0.24 | 0.70 | 71.43% | 7.60 | 1.40% |
| 6 | 33.33% | 8.14 | 0.36 | 0.50 | 33.33% | **6.87** | 0.31 | 0.52 | 83.33% | 3.05 | 0.75% |
| 7 | 71.43% | 15.03 | 0.72 | 0.54 | 71.43% | **12.88** | 0.63 | 0.59 | 71.43% | 5.02 | 0.84% |
| 8 | 50.00% | 2.56 | 0.13 | 0.70 | 50.00% | **1.54** | 0.116 | 0.72 | 100.00% | 4.50 | 0.51% |
| 9 | 50.00% | 7.97 | 0.34 | 0.58 | 50.00% | **4.58** | 0.20 | 0.65 | 50.00% | 4.43 | 0.06% |
| 10 | 100.00% | 16.64 | 0.72 | 0.65 | **100.00%** | **13.87** | 0.61 | 0.71 | 66.67% | 7.06 | 1.25% |
| Total | 55.26% | 8.21 | 0.36 | 0.61 | **63.16%** | **6.60** | 0.30 | 0.68 (**p < 0.01**) | 73.68% | 4.17 | 0.50% |

**FA/24hrs (seizure prediction):** False alarms per 24 hours, where multiple false alarms occurring within the same hour are treated as one false alarm. **FP/24hrs (seizure detection):** False positives per 24 hours, where false positives are counted for every 12-second segment, and multiple false positives within one minute period are considered as one false positive. **TW:** Time warning is calculated by total alarm time over the total recording time (for the prediction the alarm raise for one hour, while for detection alarm raise for 12 second). **Score:** The score is calculated based on the formula explained in the 4.1.7. **p:** the p value is between the 10 patients score without AURA and with AURA. **Without AURA learning:** The prediction model is pre-trained on the EPILEPSIAE dataset and is pseudo-prospectively tested using inference-only on the RPAH dataset. **With AURA learning:** The prediction model is pre-trained on the EPILEPSIAE dataset and AURA is applied to perform patient-based online training. At the same time, pseudo-prospective testing is performed on the RPAH dataset. **Detection performance:** The detection model is pre-trained on the TUH dataset and is pseudo-prospectively tested using inference-only on the RPAH dataset.

## 7 Conclusion

Human labeling of EEG data is an expensive and laborious process. Our approach to semi-supervised learning using the proposed AURA system shows a potential direction in overcoming the challenges and costs of individualized labeling to deploying patient-specific models that adapt to patient-specific bio-markers. AURA shows robustness to the use of prediction and detection models that are pre-trained on out-of-distribution data, and beyond precision medicine applications in seizure detection, AURA is a potential way to harness the multitude of unlabeled time-series clinical data that remains underutilized in deep learning. The RPAH data set is limited in the number of sessions (and therefore, ictal events) per patients which adds to the difficulty of training AURA, but we successfully demonstrated an improvement for each tested patient despite this constraint, all without the use of clinician labels. Although more experiments need to be conducted to verify the efficacy of our proposed system on wider populations and broader range of seizure types, our early demonstration shows promising utility for real-world clinical utility.

## Data Availability

The Temple University Hospital dataset is publicly available at https://www.isip.piconepress.com/projects/tuh_eeg/html/downloads.shtml. The training on the publicly available TUH dataset provides readers with sufficient level of ability to independently confirm some of the results reported. The EPILEPSIAE dataset is available at cost via http://www.epilepsiae.eu/project_outputs/european_database_on_epilepsy. The Royal Prince Alfred Hospital was used under ethics Review Board approval for our use only.

## 8 Acknowledgement

The authors would like to thank Ms. Christina Maher at the School of Biomedical Engineering, Faculty of Engineering, The University of Sydney for her contribution on our ethics process and revisions.

## Footnotes

1 See the cheap gradient principle [62].

